# The production and clinical implications of SARS-CoV-2 antibodies

**DOI:** 10.1101/2020.04.20.20065953

**Authors:** Qianfang Hu, Xiaoping Cui, Xinzhu Liu, Bin Peng, Jinyue Jiang, Xiaohui Wang, Yan Li, Wenhui Hu, Zhi Ao, Jun Duan, Xue Wang, Linxiao Zhu, Guicheng Wu, Shuliang Guo

**Author notes:** **Corresponding Authors** Shuliang Guo. PhD., Department of Respiratory and Critical Care Medicine, the First Affiliated Hospital of Chongqing Medical University, No.1, Youyi Road, Yuzhong District, Chongqing, Postal code: 400016., P. R. China., Guicheng Wu. MD., Department of Hepatopathy, Chongqing Three Gorges Central Hospital, No.165, Xincheng Road, Wanzhou District, Chongqing, Post code: 404100.P.R.China. Contributed equally (Qianfang Hu, Xiaoping Cui, Xinzhu Liu, Bin Peng). **Funding** 1. Chongqing Education Board “new coronavirus infection and prevention” emergency scientific research project (KYYJ202006). 2. Chongqing Science and Technology Bureau “new crown pneumonia epidemic emergency science and technology special” the fourth batch of projects. 3. Famous teacher project of Chongqing talent plan.

## Abstract

**Background:** Severe acute respiratory syndrome coronavirus 2(SARS-CoV-2), a novel betacoronavirus, has caused an outburst of pneumonia cases in Wuhan, China. We report the production of specific IgM and IgG antibodies after the infection of SARS-CoV-2 and its implication for the diagnosis, pathology and the course of the disease as well as the recurrence of positive nucleic acid tests after discharge.

**Methods:** Test results for SARS-CoV-2 IgM and IgG antibodies of 221 confirmed COVID-19 patients were retrospectively examined, and their clinical data were collected and analyzed based on various subgroups. SARS-CoV-2 IgM and IgG antibodies were determined with the chemiluminescence method.

**Findings:** The concentration (S/CO) of SARS-CoV-2 IgM and IgG antibodies peaked on day 19-21 after symptom onset, with a median of 17.38 (IQR 4.39-36.4) for IgM and 5.59 (IQR 0.73-13.65) for IgG. Detection rates reached highest on day 16-18 and day 19-21 for IgM and IgG, which were 73.6% and 98.6%, respectively, with significantly higher concentration of IgG in critically ill patients than in those with mild to moderate disease (P=0.027). The concentration of the antibodies on day 16-21 is not correlated with the course or outcome of the disease (Spearman r < 0.20, P > 0.05). Nasopharyngeal swabs revealed positive SARS-CoV-2 RNA in up to 52.7% of recovered patients after discharge, whose IgG proved to be significantly lower than that of those with negative RNA results (P = 0.009). IgG and IgM were tested twice within 14 days after discharge with a 7-day interval, and the second testing of these antibodies displayed a decrease in concentration of 21.2% (IQR, 11.2%,34.48%) for IgG and 23.05% (IQR, −27.96%,46.13%) for IgM, without statistical significance between the patients with re-detectable positive RNA results and those with negative RNA results after discharge. However, those with positive results experienced a count decrease in lymphocyte subsets.

**Interpretation:** The concentration of SARS-CoV-2 IgM and IgG antibodies peaked on day 19-21 after symptom onset, and antibody testing on day 16-21 is associated with increased detection rates, but the antibody concentration does not affect the course and outcome of the infection. Recovering patients with re-detectable positive SARS-CoV-2 RNA displayed lower concentration of IgG, but the downward trend of IgG during recovery indicated its limited duration of protection, and the protective effect of IgG remains to be investigated.

**Funding:** Chongqing Education Board, Chongqing Science and Technology Bureau, Famous teacher project of Chongqing talent plan

## Introduction

Corona virus disease 2019 (COVID-19), which is the infection of severe acute respiratory syndrome coronavirus 2 (SARS-CoV-2), has spread globally since its first appearance in Wuhan, China in December, 2019,^1^ accounting for 638146 cases and 30039 deaths in 203 countries, areas or territories.^2^ SARS-CoV-2 is the seventh coronavirus known to infect humans; SARS-CoV, MERS-CoV and SARS-CoV-2 can cause severe disease, whereas HKU1, NL63, OC43 and 229E are associated with mild symptoms.^3^ The infection of SARS-CoV-2 provokes immunological defense and production of specific antibodies, among which IgM antibodies are indicators of current or recent infection as the earliest to develop after exposure to the pathogen, while IgG antibodies, being the most common antibodies of the immunological response, indicates recovery of the disease or past infection. Therefore, the testing of SARS-CoV-2 IgG and IgM antibodies facilitates not only the diagnosis of COVID-19, but the evaluation of infection status as well. In fact, the testing of SARS-CoV-2 IgG and IgM antibodies has been investigated with regard to its value in early diagnosis of the disease and has been adopted as one of the diagnosis criteria in the Chinese guideline.^4,5^ However, the production of the antibodies and their protective effect, implication on the outcome and association with re-detectable positive RNA remain to be clarified. This work represents a retrospective analysis of 211 confirmed COVID-19 patients in Chongqing, China with their antibody testing to throw light on the development of SARS-CoV-2 IgG and IgM antibodies and their association with the course and outcome of the disease and re-detectable positive RNA testing after discharge.

## Methods

### Patients

The study included 211 confirmed COVID-19 patients in Chongqing Three Gorges Central Hospital from January 23^rd^ to March 3^rd^, 2020, among which were 181 mild and moderate cases (the mild group) and 40 severe and critical cases (the severe group). There were 86 female and 135 male patients, with an average age of 47·8 (47·8±15·1) years. This study was approved by the Medical Ethical Committee of the First Affiliated Hospital of Chongqing Medical University (approval number 20200601). Due to the special reasons of the epidemic, the patients’ informed consent was not obtained.

### Data collection

Clinical data including epidemiology, clinical manifestations, laboratory, course, outcome and follow-up were collected. Testing of SARS-CoV-2 IgG and IgM antibodies was performed every 3 days post symptom onset (PSO) and the results were analyzed for production, detection rates and difference of concentration between the mild group and the severe group. The 145 patients with initial antibody testing results upon admission were divided according to positive and negative IgM and positive and negative IgG, and the following information and their association with antibody production were examined: age, the length between antibody detection and symptom onset, severity of the disease, comorbidity, fever at symptom onset, count of white blood cells, neutrophils, lymphocytes, T and B lymphocytes, CD4 and CD8 lymphocytes, natural killer cells and CD4/CD8 ratio, procaicitonin, interleukin-6, tumor necrosis factor-α, γ-interferon within 3 days after admission and pulmonary inflammation index (PII) value calculated according to Jiong Wu *et al*.^6^ The relation of antibody concentration with the duration of positive virus detection (the duration between first and last positive nucleic acid testing), duration of fever (the duration between first and last detection of fever as defined by temperature > 37·3°C), length hospital stay, the progression of PII (PII value on day 7-10 after admission minus that within 3 days after admission) and outcome of the disease was investigated with the antibody testing result of 78 patients on day 16-21 after symptom onset. The predictive value of SARS-CoV-2 IgG and IgM antibody concentration for re-detectable positive nucleic acid testing was investigated with antibody testing results of 74 recovered patients within 7 days after discharge. The difference in antibody concentration between the patients with and without re-detectable positive nucleic acid was studied with two repeated antibody tests performed on day 1-7 and day 8-14 with a 7-day interval in 40 patients, and the lymphocyte subsets of the 20 patients with re-detectable positive nucleic acid were examined.

### Diagnosis, evaluation and discharge criteria

Diagnosis and discharge decisions were made according to *Guidelines for the Diagnosis and Treatment of Novel Coronavirus Infection by the National Health Commission (Trial Version 6)*. Suspected cases were subjected to reverse-transcription polymerase chain reaction (RT-PCR), and documented cases were categorized into mild, moderate, severe and critical types by clinical manifestations. Discharge criteria included: 1) normal temperature lasting over 3 days; 2) significant improvement of respiratory symptoms; 3) significant improvement of chest radiology; 4) negative nucleic acid testing in two consecutive respiratory specimens collected with an interval of at least 1 day.

### Detection of the antibodies

Testing of SARS-CoV-2 IgG and IgM was performed with serum samples which had been kept at 56 °C for 30 min, using a CFDA approved Magnetic Chemiluminescence Enzyme Immunoassay (MCLIA) kit supplied by Bioscience Co., Ltd (Chongqing, China) following the manufacturer’s instructions. The MCLIA for IgG or IgM detection was developed based on the double-antibodies sandwich immunoassay. The recombinant antigens containing the nucleoprotein and a peptide from the SARS-CoV-2 spike protein were conjugated with fluorescein isothiocyanate (FITC) and immobilized on the anti-FITC antibody-conjugated magnetic particles. Alkaline phosphatase-conjugated human IgG/IgM antibody was used as the detection antibody. The tests were conducted on an automated magnetic chemiluminescence analyzer (Axceed 260, Bioscience, China) according to the manufacturer’s instructions. Antibody levels were expressed as the ratio of the chemiluminescence signal to the cutoff value (S/CO). S/CO < 1·0 was designated as negative and otherwise was positive.

### Statistical analysis

Statistical analysis was performed using the SAS 9·4 software. The dynamic changes of the antibodies were analyzed with the logarithm of the IgG and IgM value using a generalized linear mixed model and mapped by locally weighted polynomial regression. Count data were presented as frequency with percentage and compared using χ^2^ test or Fisher’s exact test. Measurement data were presented as mean with standard deviation or median with interquartile range and comparison was made using Student’s t test or Wilcoxon rank sum test according to the normality of the data. Correlation between variables were examined with Spearman’s rank correlation.

## Results

### Trends in the development of SARS-CoV-2 IgG and IgM antibodies

The analysis included 993 test results of 221 patients, whose characteristics were listed in Supplementary Table S1. The patients in the severe group were older than those in the mild group and the difference is statistically significant (P=0·0001). Patients with diabetes, cardiovascular diseases and chronic obstructive pulmonary disease were more prone to severe and critical disease (P=0·0007, P=0·01 and P=0·004). Analysis of SARS -CoV-2 IgG and IgM antibodies examined every 3 days revealed increasing antibody levels which peaked on day 19-21, with a median concentration (S/CO) of 17·38 (IQR 4·39, 36·4) and 5.59 (IQR 0·73, 13·65), respectively, and then IgM decreased gradually while IgG remains at the high level (Figure 1). As early diagnostic indicators, SARS-CoV-2 IgM and IgG displayed detection rates of 73·58% (39/53) on day 13 -15 and 97·87% (47/48) on day 16 -18, respectively (Figure 2 and Supplementary Table S2). The trends in IgG and IgM of the patients in the severe and mild groups were shown in Figure 4 and 5. The IgG level was significantly higher in the severe group than in the mild group (F=4·28, P=0·039), while the IgM level in the two groups was not significantly different (F=0·32, P=0·57).

**Figure 1.**
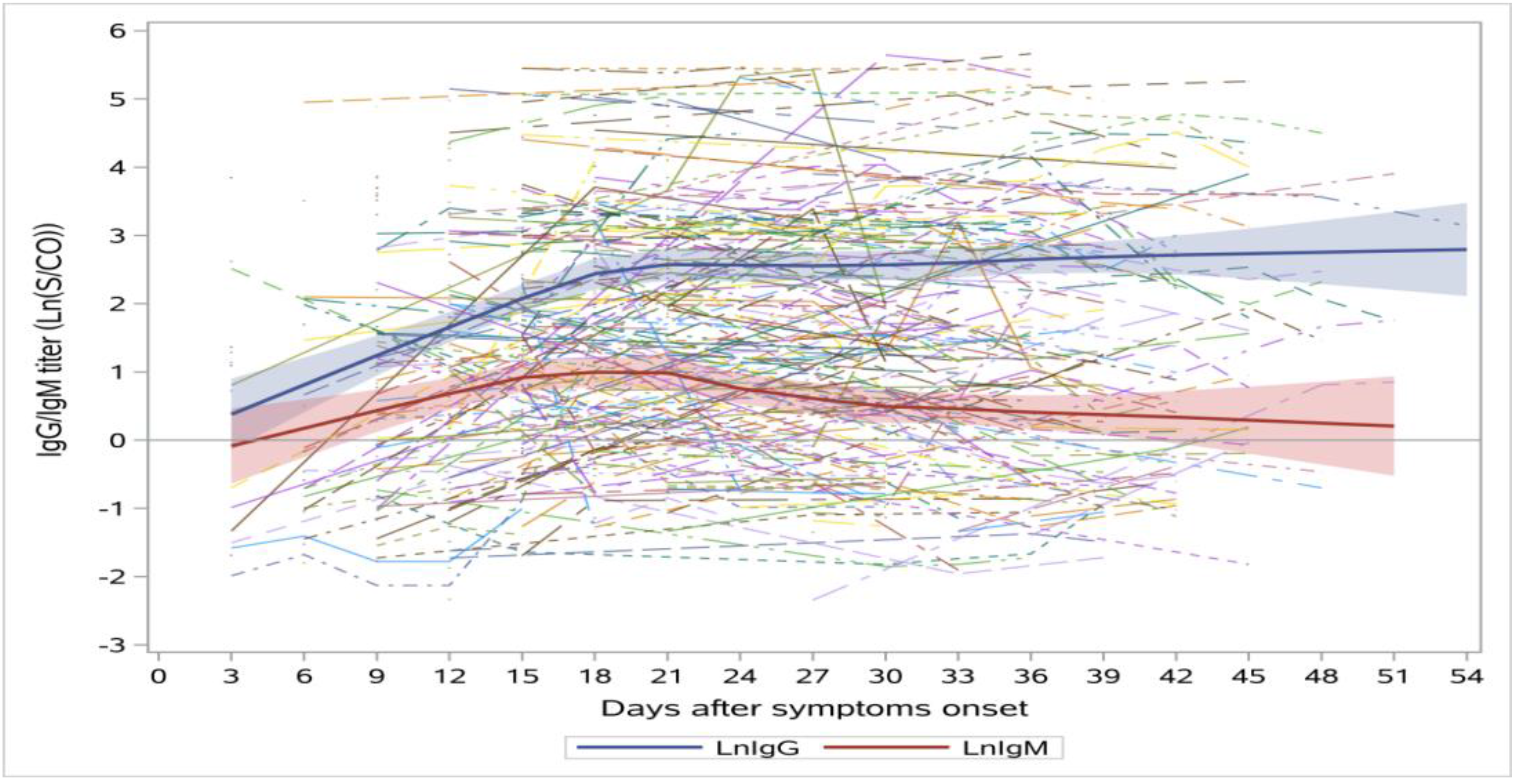
Trends in the concentration of SARS-CoV-2 IgG and IgM antibodies with time after symptom onset.

**Figure 2.**
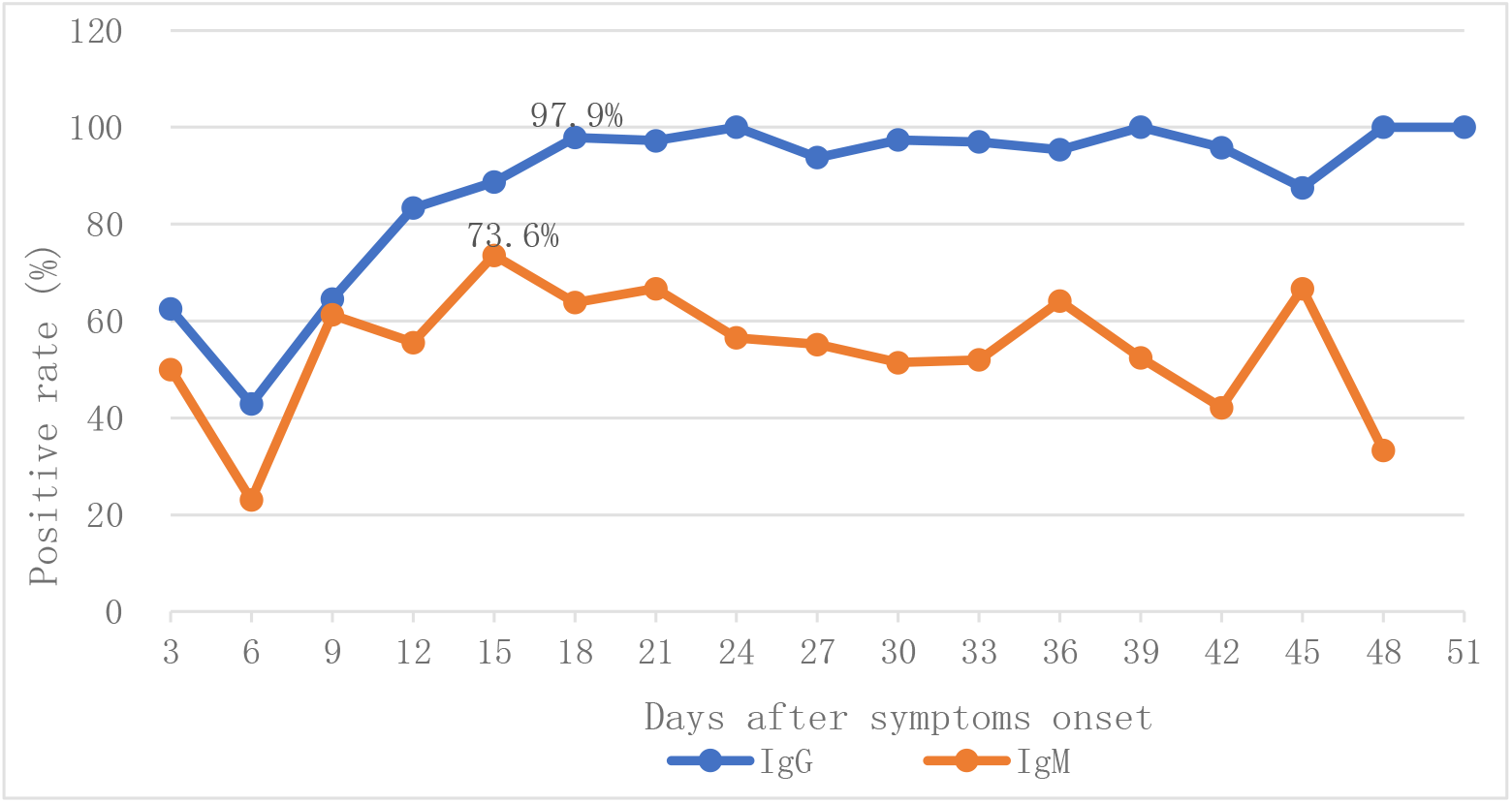
Trends in the detection rates of SARS-CoV-2 IgG and IgM antibodies with time after symptom onset.

**Figure 3.**
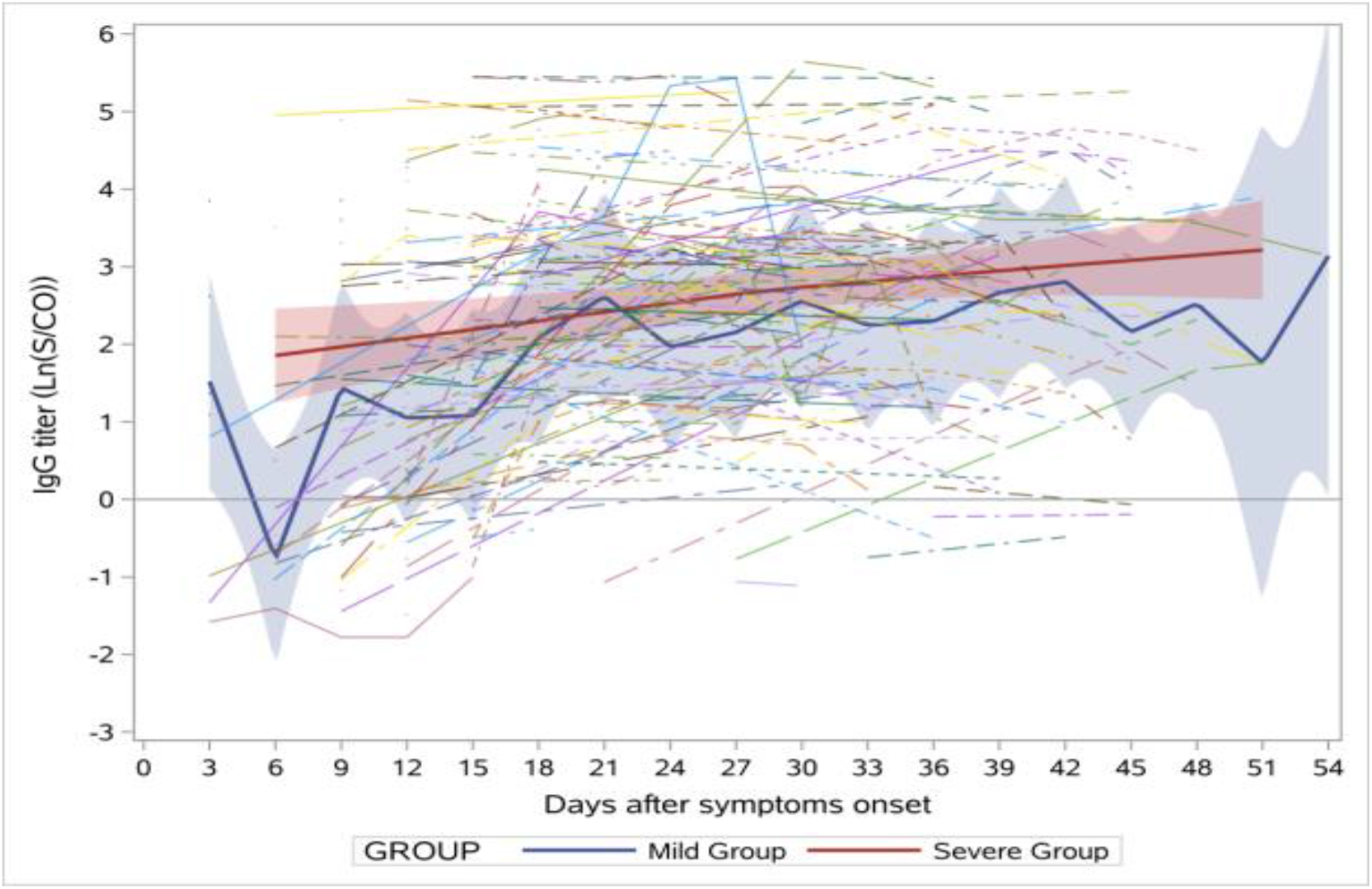
Trends in SARS-CoV-2 IgG in the mild and the severe groups.

**Figure 4.**
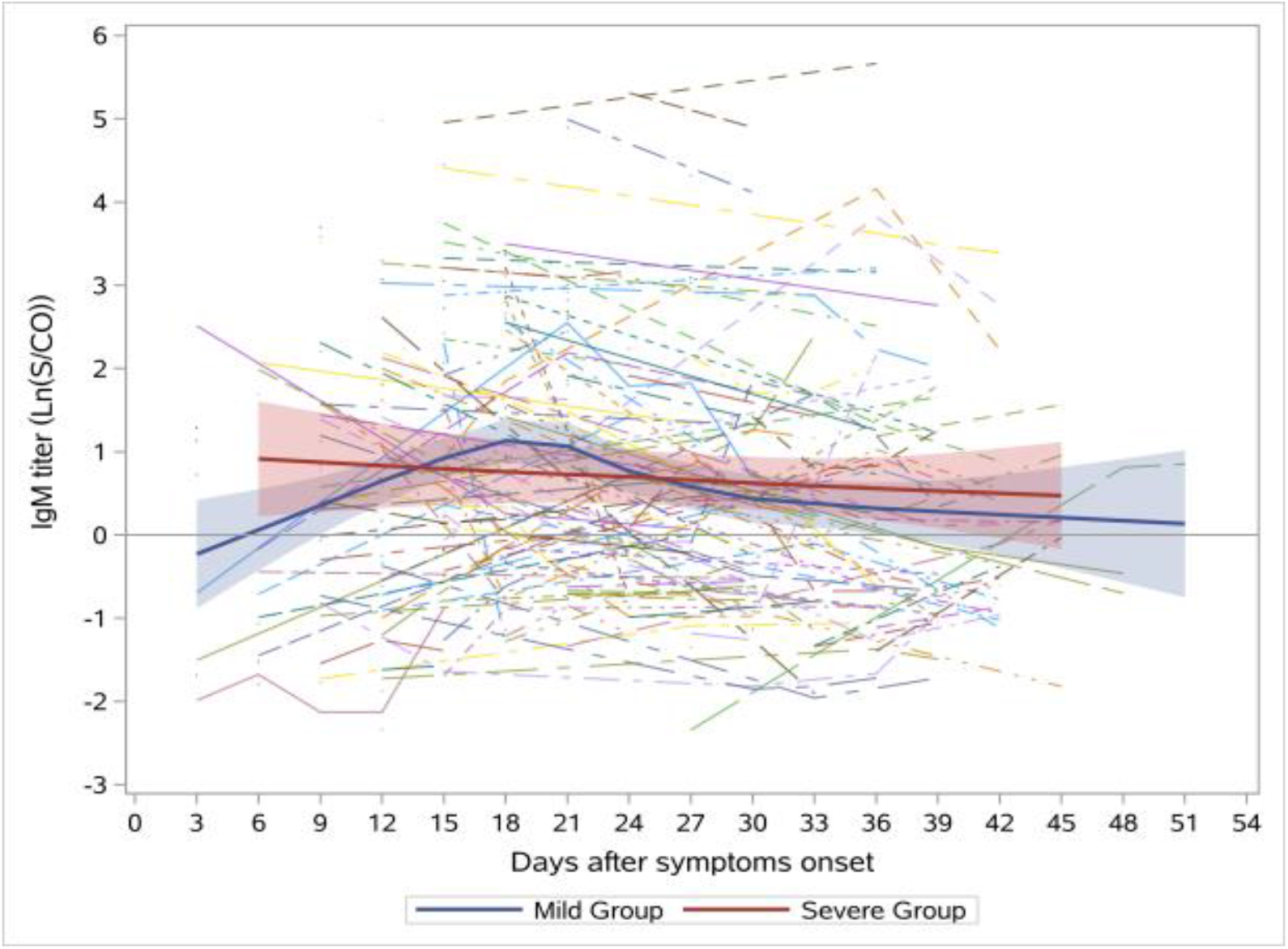
Trends in SARS-CoV-2 IgM in the mild and the severe groups.

### Factors affecting the development of SARS-CoV-2 IgG and IgM

Analysis of the initial antibody testing results upon admission of 145 confirmed COVID-19 patients revealed no statistically significant difference between the positive and negative IgM groups and between the positive and negative IgG groups regarding age, the length between antibody detection and symptom onset, severity of the disease, comorbidity, fever at onset, count of white blood cells, neutrophils and platelets within 3 days after admission, interleukin-6, tumor necrosis factor-α, γ-interferon, count of lymphocytes, T and B lymphocytes, CD4 and CD8 lymphocytes and natural killer cells and CD4/CD8 ratio. However, the PII value was significantly higher in the negative IgM group than in the positive IgM group (P=0·007, Supplementary Table S3).

### The association of SARS-CoV-2 IgG and IgM antibody concentration on day 16-21 after symptom onset with the outcome of the disease

This study revealed high concentrations of SARS-CoV-2 IgG and IgM on day 16-21 after symptom onset, and therefore the association of antibody concentration with the duration of positive virus detection, duration of fever, length hospital stay and the changes in PII within 7 days after admission was investigated. The IgG level on day 16-21 after symptom onset was not correlated with the length hospital stay (Spearman r = 0·16, P=0·18), the duration of positive virus detection (Spearman r = 0·15, P=0·20), the duration of fever (Spearman r = −0·1, P=0·39) or the changes in PII (Spearman r = 0·07, P=0·50). The IgM level on day 16-21 after symptom onset was not correlated with the length hospital stay (Spearman r = 0·03, P=0·79), the duration of positive virus detection (Sp earman r = 0·05, P=0·69), the duration of fever (Spearman r = −0·03, P=0·76) or the changes in PII (Spearman r = 0·20, P=0·11. Similarly, there were no correlation between the outcome (exacerbation or improvement) and the IgG (P=0·126) or IgM (P=0·172) lev el.

### The association of SARS-CoV-2 antibodies with re-detectable positive virus nucleic acid in recovered patients after discharge

There were 74 recovered patients who met the discharge criteria and were discharged to isolation with medical observation for 14 days, and up to 39 (52·7%) of them presented with re-detectable positive virus nucleic acid during this period. These patients had significantly lower IgG concentration within 7 days after discharge (P=0·009), but the difference in IgM concentration wa s not significant (P=0·06). The decrease of SARS-CoV-2 IgG and IgM antibodies in 40 recovered patients between two tests within 14 days after discharge with a 7-day interval was not statistically significant (P=0·16 and P=0·265, respectively), but the decrease of IgG and IgM each reached 21·2% (IQR, 11·2%, 34·48%) and 23·05% (IQR, −27·96%, 46·13%) (Table 1 & 2). In addition, decrease was seen in the count of total lymphocytes, total T lymphocytes, CD4 lymphocytes, CD8 lymphocytes, total B lymphocytes and natural killer lymphocytes to various extents, accounting for 90%, 55%, 65%, 25%, 55% and 20% of the 20 patients with re-detectable positive virus nucleic acid when readmitted, respectively (Supplementary Table S4)

**Table 1.** SARS-CoV-2 IgG and IgM antibodies in recovered patients within 7 days after discharge.

|  | Total (n=74) | Negative Cases (n=35) | Positive Cases (n=39) | P value |
| --- | --- | --- | --- | --- |
| Age, years | 47.5 ± 15.7 | 45.2 ± 15.6 | 49.6 ± 15.7 | 0.24 |
| Sex |  |  |  |  |
| Female | 30 (39.2%) | 16 (33.3%) | 14 (44.4%) | 0.74 |
| Male | 44 (60.8%) | 19 (66.7%) | 25 (55.6%) |  |
| IgG, S/CO | 15.76 (3.72, 28.56) | 20.19 (7.75, 70.50) | 8.94 (2.68, 21.4) | 0.009 |
| IgM, S/CO | 1.18 (0.53, 3.82) | 1.39 (0.76, 3.94) | 0.90 (0.40, 3.79) | 0.06 |

Data are median (IQR) or n (%). Positive cases, recovered COVID-19 patients with re-detectable SARS-CoV-2 nucleic acid of nasopharyngeal swabs within 14 days after discharge; negative cases, recovered COVID-19 patients with negative SARS-CoV-2 nucleic acid of nasopharyngeal swabs within 14 days after discharge.

**Table 2.** Decrease of SARS-CoV-2 IgG and IgM antibodies in recovered patients between two tests within 14 days after discharge with a 7-day interval.

|  | Total (n=40) | Negative Cases (n=19) | Positive Cases (n=21) | P value |
| --- | --- | --- | --- | --- |
| Age, years | 47.3 ± 14.7 | 45.6 ± 16.7 | 49.1 ± 16.7 | 0.459 |
| Sex |  |  |  | 0.100 |
| Female | 21 (52.5%) | 10 (52.6%) | 11 (52.4%) |  |
| Male | 19 (47.5%) | 10 (52.6%) | 9 (42.9%) |  |
| The difference of IgG*, S/CO | 3.47 (0.35, 5.55) | 4.95 (1.1, 5.83) | 1.76 (-0.24, 4.54) | 0.16 |
| The difference of IgM*, S/CO | 0.13 (-0.45, 0.8) | 0.44 (0.27, 1.01) | 0.04 (-0.4, 0.2) | 0.27 |
| The decrease in IgG#, (%) | 21.2 (11.2, 34.48) | 22.4 (14.5, 34.48) | 17.78 (0.28, 34.48) | 0.32 |
| The decrease in IgM#, (%) | 23.05 (-27.96, 46.13) | 30.64 (1.62, 48.8) | 1.35 (-29.27, 30.36) | 0.21 |

Data are median (IQR) or n (%). \* The difference of two antibody tests within 14 days after discharge with a 7-day interval. # The decrease of antibody level detected within 14 days after discharge with a 7-day interval, as calculated by (difference of the results of the two antibody tests/result of the first antibody test) x100.
Positive cases, recovered COVID-19 patients with re-detectable SARS-CoV-2 nucleic acid of nasopharyngeal swabs within 14 days after discharge; negative cases, recovered COVID-19 patients with negative SARS-CoV-2 nucleic acid of nasopharyngeal swabs within 14 days after discharge.

## Discussion

As the epidemic escalates, antibody testing has been included in the diagnostic procedure according to Chinese guidelines, and there are several CFDA-approved diagnostic kits available for detection of SARS-CoV-2 IgG and IgM, which have shown a sensitivity of 70·24% and 9 6·10% for IgG and IgM, respectively.^7^ This study demonstrated an upward trend of detection rate of SARS-CoV-2 IgG and IgM after symptom onset. The detection rate of IgM peaked at 73·6% on day 13 -15 and later decreased gradually, while that of IgG reached 97·8% on day 16 -18 and then remained at the high level. However, there were 2 patients who did not develop either IgG or IgM antibody during the whole course of disease. Therefore, SARS-CoV-2 IgG and IgM antibodies testing should be combined with RT-PCR as an early diagnosis method,^8^ and negative antibody testing could not rule out the disease. The chronological mapping of the level of SARS-CoV-2 IgG and IgM antibodies revealed a peak of both antibodies on day 19-21, after which IgM gradually decreased while IgG remained at a high level, which was consistent with the development of IgM. It has been reported that SARS-CoV and SARS-CoV-2 IgG and IgM peak on the 3^rd^ week, after which IgM persistently decrease while IgG remains at a high level,^9,10^ but the analysis of SARS-CoV-2 IgG and IgM tests in 40 recovered patients after discharge with a 7-day interval demonstrated a median decrease of 21·2% in IgG regardless of re-detectable positive nucleic acid, indicating instant decrease of IgG after recovery and, therefore, that long-term protection provided by IgG requires further study, which may be helpful for vaccine development and raises questions on the timing of serum collection of recovered patients for treatment.

The analysis of possible influencing factors of antibody development did not reveal statistical significance except for PII value, but the first part of the study indicated an increasing trend in the detection rate of the antibodies which peaked on day 13-15 for IgG and on day 16-18 for IgM. The first antibody test in this study was conducted on day 13 of symptom onset (IQR 11^th^ −16^th^ day), possibly due to the period of research, development and approval of the antibody detection kits as COVID-19 was caused by a novel coronavirus. However, a report by Long *et al*. found that the median time until seroconversion of SARS-CoV-2 IgG and IgM was 13 days,^12^ so the fact that antibody tests were conducted after day 13 does not weaken the findings of this study. Nonetheless, the higher PII value in the negative IgM group in comparison to the positive IgM group indicated severer respiratory symptoms in patients with slower acute humoral immune response.

Until the validation of specific therapy for SARS-CoV-2, host immune system remains to be the most effective defense against the disease. This study found no correlation between IgG and IgM concentration and viral shedding, course and outcome of the disease through a retrospective analysis without the knowledge of SARS-CoV-2 viral load. Similarly, the study by Corman *et al*. did not find evidence for inhibition of MERS-CoV by neutralizing antibodies.^11^ The kit in this study mainly detected antibodies against nucleoprotein and spike protein due to the use of recombinant antigens containing the nucleoprotein and a peptide from spike protein of SARS-CoV-2,^12^ and the IgG antibody against spike protein has been proved to induce severe acute lung injury in Chinese macaques through alterations in inflammation response as reported by Liu *et al*.^*13*^ Similar lung injury caused by SARS-CoV vaccines has been observed in other animal models, such as mice and African green monkeys, ^14-17^ and higher IgG concentration has been detected in severe and critical patients with COVID-19. Two other studies of 222 and 173 COVID-19 patients also revealed worse prognosis associated with higher IgG concentration.^18,19^ Therefore, the protective effect of IgG and its association with the deterioration of lung injury remain to be explored with further researches, and these findings have also raised questions on validity of the practice of treating the disease with the serum of recovering patients despite encouraging results of clinical improvement in all 5 critically ill patients after transfusion of convalescent plasma containing neutralizing antibodies in a preliminary trial.^20^

This study demonstrated a rate of re-detectable positive nucleic acid as high as 52.7% (39/74) despite strict adherence to domestic guidelines on discharge criteria, which may be attributed to the fact that SARS-CoV-2 mainly resides in the lower respiratory system.^21,22^ In addition, possible deviation in specimen collection may lead to false negativity of nucleic testing and consequently inappropriate discharge decisions,^23,24^ resulting in recurrence and even deterioration of the disease. The 20 patients with re-detectable positive nucleic acid displayed decline in lymphocyte subsets on the second admission, which was similar to the changes of initial infection.^25-29^ These patients with SARS-CoV-2 nucleic acid detected in nasopharyngeal swabs within 14 days after discharge had significantly lower IgG concentration compared to those with negative nucleic acid after discharge, but the difference in IgM concentration was not significant. Meanwhile, the changes in IgG and IgM tests with a 7-day interval did not display significant differences. Therefore, repeated nucleic acid testing should be conducted using specimens collected at multiple sites before discharge,^30^ and the level of IgG may contribute to the discharge criteria. The results also suggested the possibility that IgG not only serves as a protective antibody, but also bears the potential to deteriorate the injury of multiple organs, which, if proven, will raise concern on treatment with serum antibody of recovered patients, and may indicate IgG or its immune complex as indicators to evaluate organ injury and disease severity.

## Data Availability

With the permission of the corresponding authors, we can provide participant data without names and identifiers, but not the study protocol, statistical analysis plan, or informed consent form. Data can be provided after the Article is published. Once the data can be made public, the research team will provide an email address for communication. The corresponding authors have the right to decide whether to share the data or not based on the research objectives and plan provided.

## Contributors

QF H, SL G, made substantial contributions to the study concept and design. QF H, XZ L,was in charge of the manuscript draft. WH H, X W and LX Z, Z A collecting and confirming data accuracy. JY J,J D, applied for the ethical approval, B P,QF H participated in statistical analysis. XP C,QF H were in charge of the laboratory tasks, including sample processing, detection, data acquisition and interpretation. Y L,GC W were the clinical expert in charge of the treatment of the patients. SL G,GC W,made substantial revisions to the manuscript.All authors contributed to data acquisition, data analysis, or data interpretation, and reviewed and approved the final version.

## Acknowledgments

This work is funded by Chongqing Education Board “new coronavirus infection and prevention” emergency scientific research project (KYYJ202006). Chongqing Science and Technology Bureau “new crown pneumonia epidemic emergency science and technology special” the fourth batch of projects. Famous teacher project of Chongqing talent plan We acknowledge all health-care workers involved in the diagnosis and treatment of patients in Wuhan; we thank the Chongqing Three Gorges Central Hospital for coordinating data collection for patients with SARS-CoV-2 infection, and we thank Prof Xinquan Long for interpretation of results.

